# Quality of systematic reviews on physiotherapy interventions for musculoskeletal disorders is critically low: a meta-epidemiological study

**DOI:** 10.1101/2023.11.20.23298761

**Authors:** Nicola Ferri, Elisa Ravizzotti, Alessandro Bracci, Giulia Carreras, Paolo Pillastrini, Mauro Di Bari

## Abstract

**Question:** How good is the quality of systematic reviews on the effectiveness of physiotherapy for musculoskeletal conditions? Are there any factors associated with quality?

**Design:** This is a meta-epidemiological study on systematic reviews with meta-analysis (SR-MA) of randomised controlled trials (RCT).

**Methods:** MEDLINE, Cochrane Database of Systematic Reviews (CDSR), CINAHL, and PEDro were searched for SR-MA of RCT on physiotherapy in musculoskeletal disorders in the last ten years. Two independent researchers screened and extracted the records and analysed the full-texts. The quality of SR-MA was quantified with AMSTAR-2 tool on a sample of 100 studies, randomly selected from the records retrieved. Disagreements were solved by consensus.

**Results:** The number of eligible publications increased over the past ten years. However, the methodological quality was critically low in as many as 90% of the studies retrieved and did not increase with time. The last author’s H-index was the only quality predictor among the variables analysed.

**Conclusion:** The methodological quality of the SR-MA of RCT is unacceptably low. Given the frequent application of physiotherapy in musculoskeletal disorders, there is an urgent need to improve secondary research by adopting more rigorous methods.

**Registration:** Open Science Framework (https://osf.io/bc8zw/)

## INTRODUCTION

Systematic reviews with meta-analysis (SR-MA) are essential to Evidence-Based Medicine, as they are considered the best synthesis of intervention studies – in particular, of randomised clinical trials (RCT) – and a fundamental source of evidence for guideline developers and policymakers.^1^ The number of this type of studies has been dramatically increasing in recent years, up to an estimate of 80 new SR-MA published daily.^2^

The area of physiotherapy has clinical, social, and economic relevance within the scientific literature. It has been estimated that one-third of all the people in the world will need physiotherapy at some point in their lives.^3^ Out of the variety of conditions that can benefit from physiotherapy, the most frequent indication to physiotherapy is represented by musculoskeletal disorders, which affect 1.71 billion people (95% CI 1.68 to 1.80) across the world, with low back pain being the leading condition.^3^

Despite an increasing information overload, little is known about the overall quality of SR-MA and the impact of their results and conclusions on clinical practice. This can undermine the progress and credibility of research and be an obstacle to reducing the gap between researchers and clinicians. Recent methodological studies, analysing SR-MA quality in specific medical areas, found that quality was low or critically low in several, diverse fields, such as treatments for Alzheimer’s disease (82.4%),^4^ acupuncture (99.1%),^5^ Chinese herbal medicine (99.3%),^6^ and surgical adverse events (100%).^7^

The “Publish or Perish” paradigm is one of the elements behind quantity over quality.^8^ For this reason, there is still debate about the strengths and limitations of current bibliometrics in representing the quality and impact of scientific research in academics.^9–12^ We have recently conducted a survey confirming that more than 50% of decision-makers have difficulty choosing the best evidence on a given topic: the methodological quality, the reputation of the authors and the journal, and the type of primary studies included were among the features thought to be important by respondents.^13^

The research questions were:

1. How good is the quality of SR-MA of RCT on the effectiveness of physiotherapy for musculoskeletal conditions published in the last ten years?
2. Is the quality associated with any publication factor?

## METHOD

### Design

This is an observational study in the physiotherapy evidence synthesis field. Since there are no specific guidelines for meta-epidemiological studies, and the MethodologIcal STudy reporting Checklist (MISTIC) is still under development,^14^ we followed the Cochrane Handbook for guidance in the selection and extraction processes,^15^ and the PRISMA 2020 statement for reporting, as applicable.^16^ This meta-epidemiological study was prospectively registered on the Open Science Framework (https://osf.io/bc8zw/).

### Population

Our study focuses on musculoskeletal conditions defined and categorised by the International Classification of Diseases 11th Revision (ICD-11).^17^

### Intervention

We included all types of treatment that physiotherapists in any part of the world can administer, considering the heterogeneity of norms and cultures: thus, acupuncture, dry needling, yoga, and percutaneous TENS were included, among other approaches. All studies on prevention in healthy subjects were excluded.

### Control

We included all types of comparisons, because our objective was to study the methodological quality, not to perform a synthesis of treatment effectiveness.

### Outcome measures

For the same reason, we had no interest in considering only specific outcomes. The only restriction applied was to studies broadly addressing the clinical effectiveness of physiotherapy: thus, economic studies were excluded.

### Study design

We included only SR-MA of RCT because they are considered as the best summary of the available scientific evidence, a claim which justifies their high impact on clinical practice in questions concerning treatment effectiveness. SR-MA incorporating also non-randomised intervention studies were included only if a meta-analysis of RCT was reported separately. We excluded network meta-analyses, scoping reviews, rapid reviews, and other forms of evidence synthesis different from Cochrane’s definition of SR.^15^

### Searching

We searched MEDLINE (via PubMed), Cochrane Database of Systematic Reviews (via Cochrane Library), CINAHL (via EBSCOHost), and PEDro, from December 2012 to December 2022; the search strategy included both structured terms and free texts linked by logical operators (see Appendix 1 on the eAddenda). Then, a semi-automated deduplication via EndNote^a^ was performed, followed by a manual check. One record was randomly selected if the same paper was published in multiple journals. The strategy for full-text retrieval included contacting authors of the manuscript when necessary.

### Screening

Two independent researchers selected records by Title and Abstracts, based on the inclusion and exclusion criteria as described above. The same process was repeated on the full-texts retrieved. Any disagreement was solved by consensus.

### Sampling

We randomised all the full-texts using an Excel^b^ function: a final sample of 100 SR-MA was deemed as adequate for the purposes of the study.

### Extraction

A researcher extracted data of interest (Table 1) from the SR-MA included, using a predefined Excel^b^ form. A second reviewer then checked all the extractions; conflicts were solved by consensus.

**Table 1.** Extraction data.

| *General variables* | *Authors variables* | *Journal variables* | *Study variables* |
| --- | --- | --- | --- |
| First author | Country | Journal IF<sup>c</sup> | Type of SR-MA<sup>d</sup> |
| Year of publication<sup>c</sup> | Number of authors<sup>c</sup> | Journal Q (Scimago)<sup>c</sup> | Publication policy<sup>d</sup> |
| Study title | First author h-index<sup>c</sup> | Journal Q (JCR)<sup>c</sup> | Total studies included<sup>c</sup> |
| Journal name | Last author h-index<sup>c</sup> |  | Study design included<sup>d</sup> |
|  |  |  | Results<sup>d</sup> |
|  |  |  | Conclusions<sup>d</sup> |

IF: Impact Factor, Q: Quartile, JCR: Journal Citation Report, SR-MA: Systematic Review with Meta-Analysis. c: continuous data, d: dichotomous data.

Type of SR-MA identifies Cochrane vs. non-Cochrane review. Journals’ impact factor (IF) was assessed at the time of the publication via the Clarivate Journal Citation Report and the same was done for journals’ Quartile via Scimago and Clarivate. When a journal was classified as belonging to different discipline areas, the quartile of the discipline closer to physiotherapy was considered. First and last authors’ h-indexes were assessed via Scopus. The journal’s access policy was determined by consulting the Directory of Open Access Journals, Clarivate Journal Citation Report, Scopus, the journals’ site, and the possibility of a full-text free download. The number of the studies included refers to the papers included in the SR, not only in meta-analyses. In extracting results and conclusions, only the primary outcome was considered; if more than one primary outcome was present or no primary outcome was mentioned, we extracted the first that was reported. We also considered the results favorable if a statistical significance was obtained and the conclusions favorable if a recommendation toward the intervention was given. Country refers to the corresponding author’s primary affiliation; we categorised this data into the continent for statistical analysis.

### AMSTAR-2 assessment

A Measurement Tool to Assess Systematic Reviews - version 2 (AMSTAR-2) is a critical appraisal tool for systematic reviews, consisting of 16 items on methodological domains with a yes/no and partial yes rating.^18^ Items number 2, 4, 7, 9, 11, 13, and 15 are considered as critical domains and, therefore, they are weighted more in the final quality assessment. The psychometric characteristics of AMSTAR-2 have been studied and validated, comparing it with pre-existing tools.^19^

Three researchers performed deep training following the AMSTAR-2 publication, guidance, and online educational support. A piloting test was completed before the assessments, and there was a discussion with AMSTAR-2’s authors on some judgment calls to enhance the most accurate interpretation and consistency. Then, two independent researchers assessed each SR included, and any conflict was solved by consensus. A summary score was not calculated, whereas, as recommended by Shea et al.,^18^ four levels of quality (i.e., critically low, low, moderate, and high) were identified.

### Data analysis

Data were analysed with descriptive statistics: we used mean and standard deviation for continuous data (or median and interquartile range for non-normal distribution), counts and percentages for categorical data. All the variables were grouped according to the AMSTAR-2 score, and statistical tests were performed to analyse any significant differences.

With the aim of preserving the ordinal distribution of the four AMSTAR-2 levels, an ordinal logistic regression analysis models was built to identify predictors of SR-MA quality among the variables extracted. Statistical analyses were performed with Stata 17 software^c^.

## RESULTS

The search strategy retrieved 2.067 records, of which 151 were duplicates, and 2 were retracted papers. Thus, 1914 records were screened by title and abstract, leading to 475 full texts assessed for eligibility. Finally, 395 SR-MA were included (Figure 1), of which a random sample of 100 was obtained. We reported details of the studies included (see Appendix 2 on the eAddenda), and the reasons for full-text exclusions (see Appendix 3 on the eAddenda).

**Figure 1.**
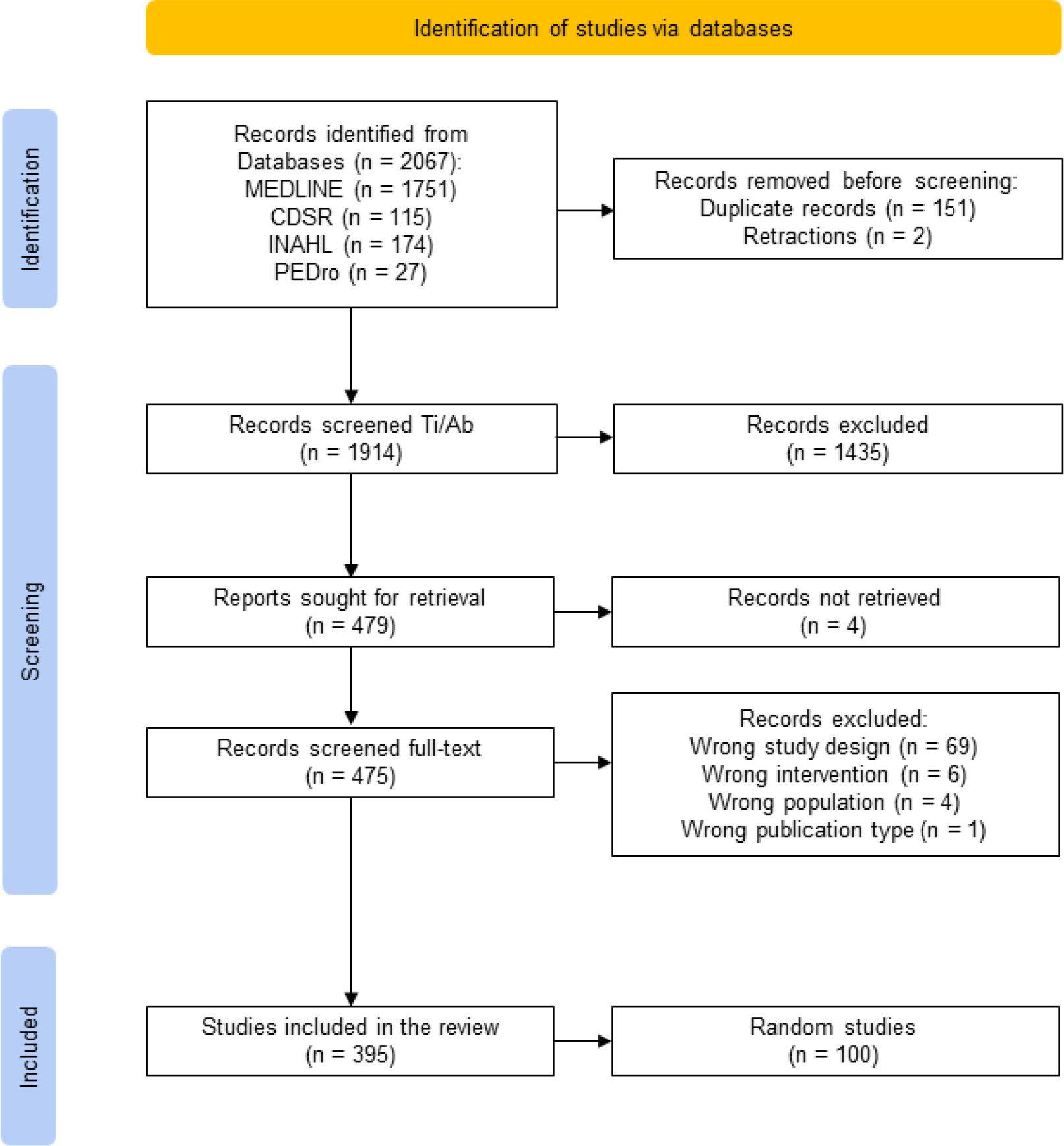
PRISMA flow diagram.

Overall, as many as 90% of the studies selected were judged as of critically low quality. Moreover, although the number of publications increased markedly over the past ten years, the methodological quality of the studies remained always markedly unsatisfactory (Table 2).

**Table 2.**
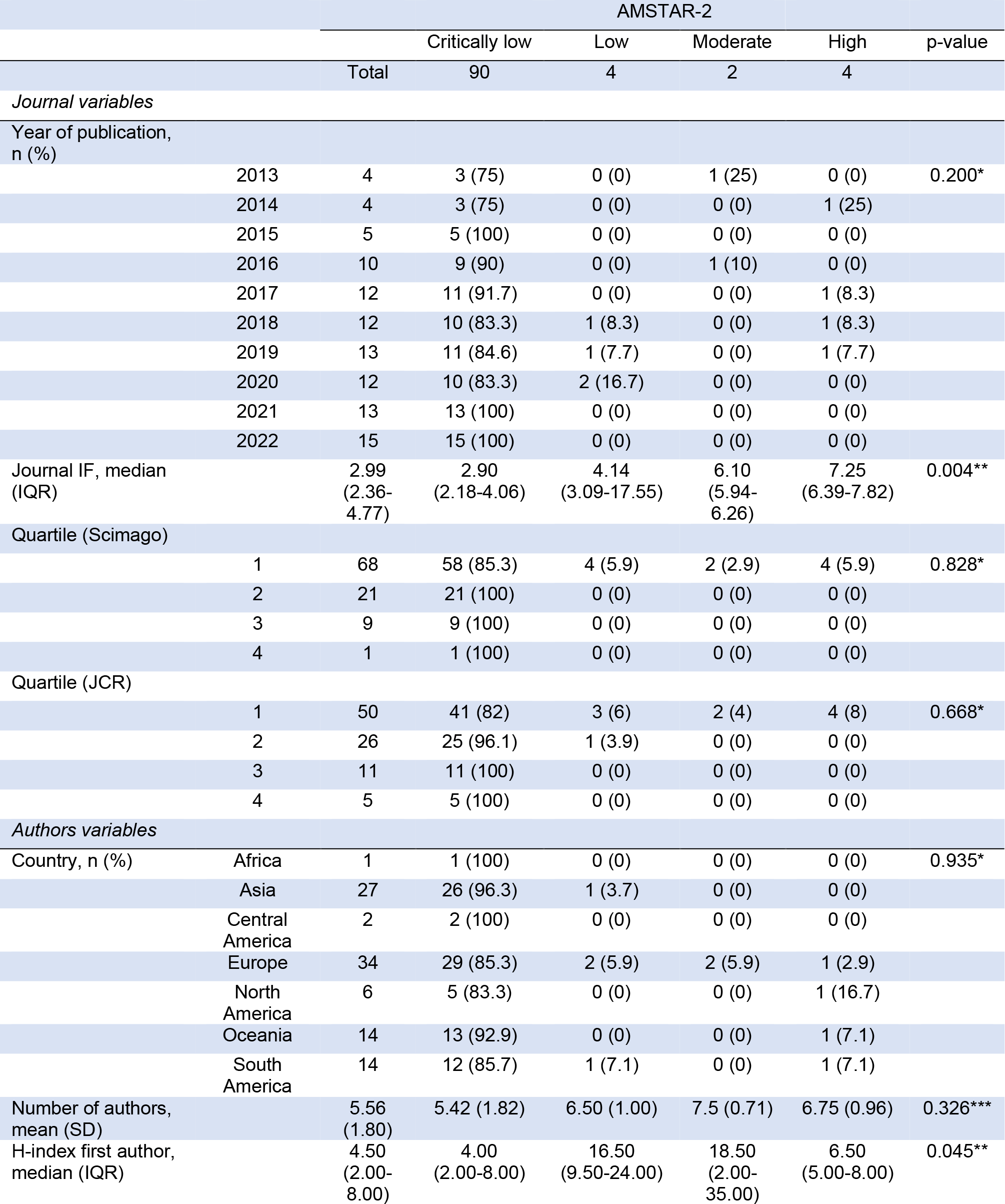
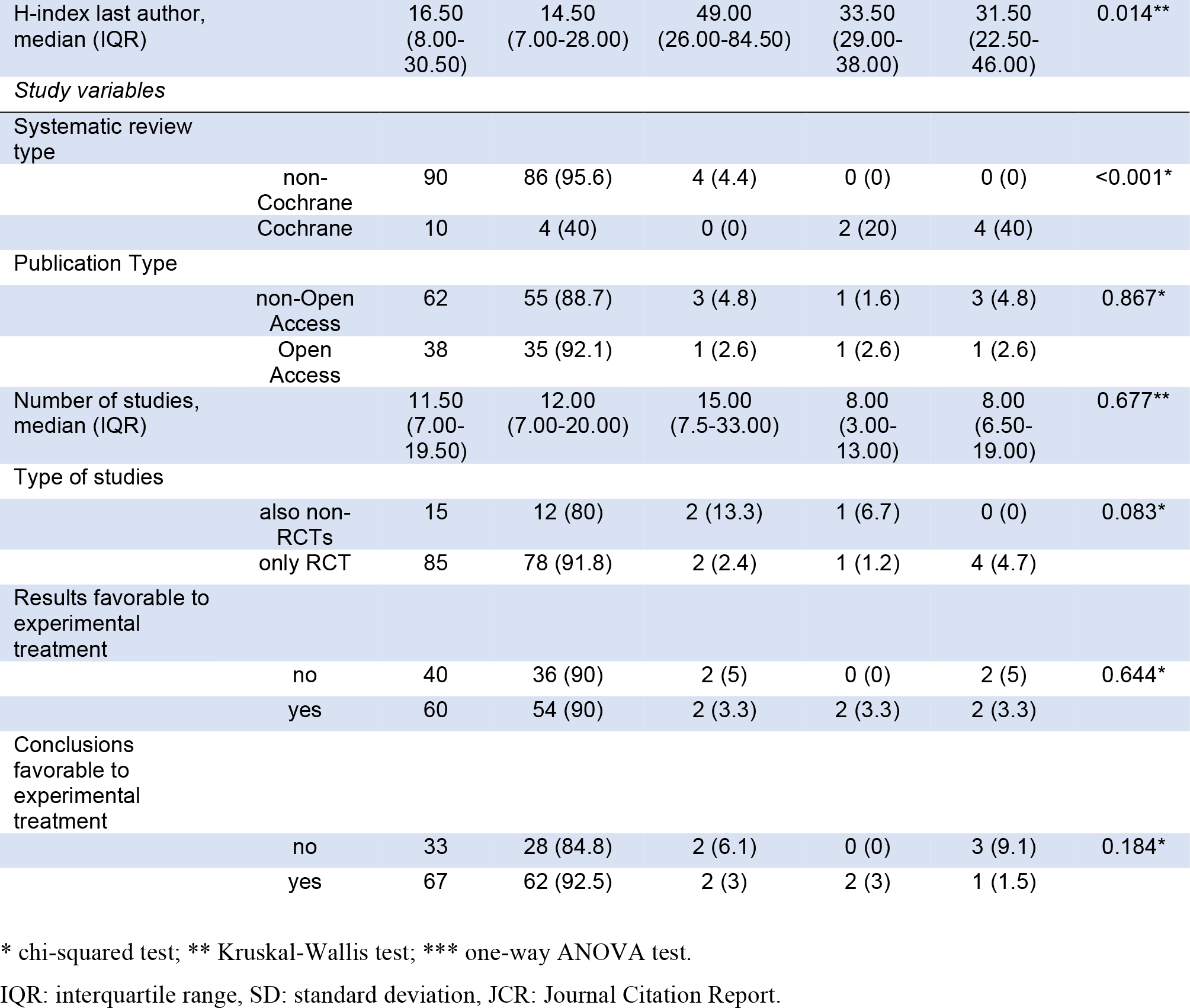
Descriptive statistics.

As shown in Table 2, every continent was represented in our sample, yet with different distributions. After exclusion of 8 journals that had no IF at the publication date, the IF ranged from

0.445 to 30.313 and the differences between the four AMSTAR-2 levels were statistically significant. The H-index of the first and last authors ranged from 0 to 39 and 1 to 100, respectively; these variables had a significantly different distribution according to the AMSTAR-2 quality scores. The only four studies rated as high quality were all Cochrane SR-MA.

Analysing the single items (Figure 2), 93% of the studies did not explain the reason for the eligibility criteria of study designs (item 3), 78% did not report the list of the studies excluded (item 7), and 90% did not check funding sources of the primary studies included (item 10).

**Figure 2.**
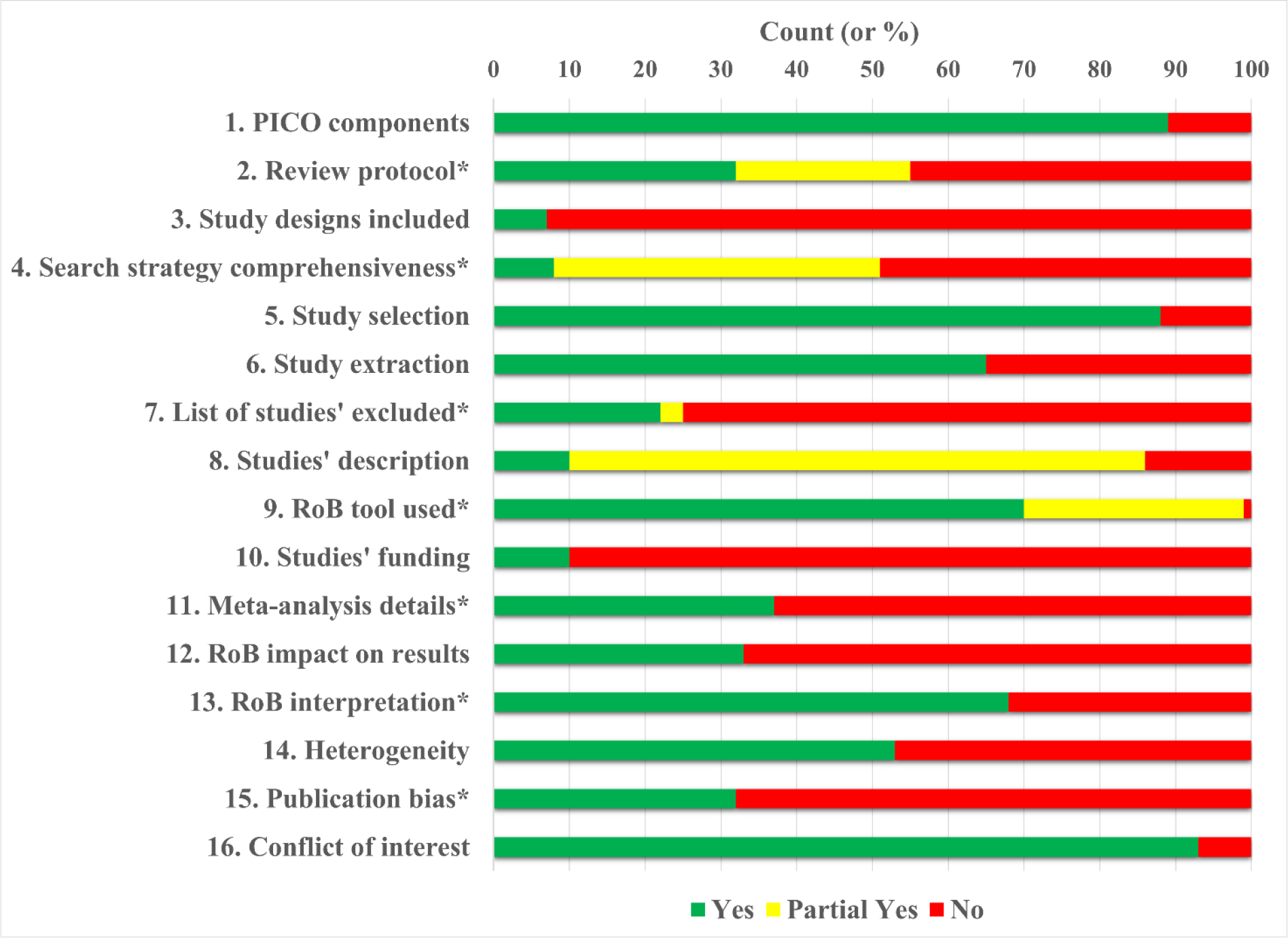
AMSTAR-2 items assessment. Asterisks identify the seven critical domains.

### Ordered logistic regression

An ordered logistic regression model for analysing AMSTAR-2 scores was built (Table 3). Covariates that satisfied the proportional odds assumption were identified, and the best fitting model in terms of Akaike Information Criterium was selected. Results are displayed as proportional odds ratios.

**Table 3.**
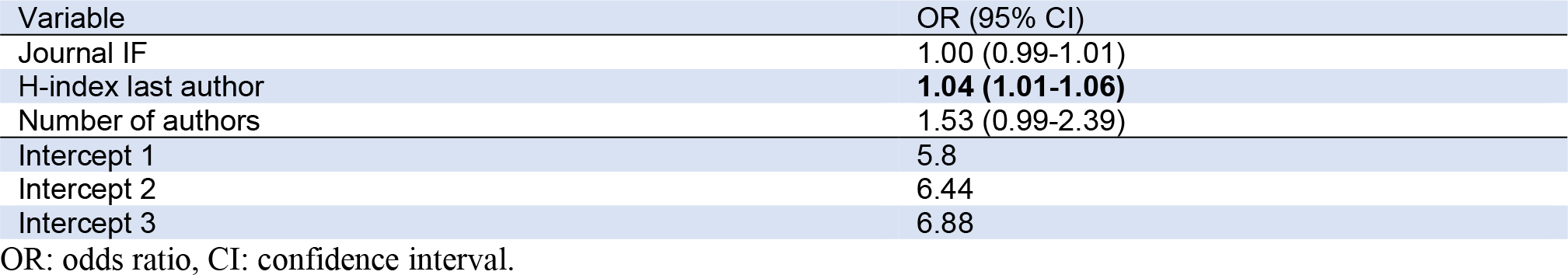
Multivariable ordinal logistic regression predicting the methodological quality of 100 systematic reviews with meta-analysis of randomised clinical trials on musculoskeletal physiotherapy.

The journal’s IF was not a predictor of quality. Conversely, the odds of a high AMSTAR-2 level versus the combined critically low, low, and moderate AMSTAR-2 level were 1.04 times greater for each unit increase in the H-index of the last author, assuming that all other variables in the model were held constant. Because of the proportional odds assumption, the same increase of 1.04 times would represent the risk of being in the combined AMSTAR-2 high and moderate categories versus the critically low and low ones. Increasing the number of authors by one resulted in 53% higher odds for high AMSTAR-2 than the other categories, with a borderline significant association.

## DISCUSSION

In this representative sample of SR-MA of RCT on musculoskeletal physiotherapy published in the last ten years, the methodological quality, assessed with a standardised and recognised tool, was critically low in as many as 90% of the studies retrieved and did not show any time-dependent increase. Of the candidate predictors, only the last author’s H-index predicted the quality level of the SR-MA.

The practice of healthcare professions implies an obligation of means toward the patient, which, in turn, translates into acting with prudence and diligence. To achieve this, both for clinical and legal purposes healthcare providers should mandatorily apply the best scientific evidences available. Yet, what if the evidence is of low quality?

This meta-research strengthens the findings of Riley et al.20, who reported low quality on a sample of 24 SRs on a similar topic. We suggest being cautious about the evidence synthesis quality, which could be a critical issue in implementing Evidence-Based Medicine in musculoskeletal physiotherapy, both in choosing the best evidence and for the risk of not properly considering the quality of conduction during knowledge translation.

Sometimes, assessing methodological quality is challenging, because it also depends on the quality of reporting; our difficulty is supported by a recent study that found a significant association between the quality of reporting and the risk of bias of SR.^21^ In our study, these challenges were overcome thanks to the extensive training and piloting preliminary conducted and the consensus-based decision-making process. Retrospectively, our choices were consistent with the latest published recommendations.^22^ We also found a tendency for a floor effect of the AMSTAR-2 tool, as described in a previous research:^23^ thus, one might wonder whether indeed the SR-MA were of low quality or the instrument was too rigid and demanding. However, a high standard is definitively desirable and, after all, we did find some high-quality reviews, a finding that clearly indicate that such levels of methodological quality can be achieved. Thus, we argue that methodological improvement is a priority.

The H-index of the last author was the only variable with a predictive value for quality. This finding is consistent with the last author being usually a senior researcher, responsible for the choice and training of collaborators and for the overall scientific architecture of the study. In a way, it also represents a small comfort on the goodness of the index, knowing all its recent limitations and criticalities within academia.^9-12^

Main strengths of this study are the comprehensive and rigorous selection of the studies to analyse, the application of a recognised, standardised tool for quality assessment, and the use of a statistical approach that accurately reflects the ordinal nature of the outcome variable. At the same time, some limitations must be acknowledged. First, in the expectation of a possible time-dependent trend in methodological quality, we included a broad and representative number of studies published in the last ten years: however, the AMSTAR-2 tool was published only in 2017, more recently than the oldest studies we selected. Nevertheless, the tool was designed to assess quality, not as guidance to conduction of a SR; moreover, a previous version of AMSTAR and the Cochrane Handbook were both already available before 2017.^24,25^ Year of publication trended to be negatively correlated with quality, although this variable could not be included in the multivariable ordinal model due to a violation of the assumption of proportionality. Second, to optimise resources, we did not analyse the entire sample of 395 SR-MA, but only a share of them. However, because the sample was drawn at random and included more than a quarter of the studies retrieved, we are confident of its representativeness and, consequently, of the validity of our results, which, indeed, are well consistent with previous studies. Last, the open-access variable was attributed not to the journal, but rather to each specific article, considering the free access to it in the year of publication. We realise that the dichotomisation open-access yes/no does not fully capture the heterogeneity of publication policies and may result in less sensitive data for this variable.

Evidence-based medicine involves considering patient expectations and seeking the best scientific evidence to support clinical reasoning and decision-making. This research alerts us to the very low quality of SR-MA on treatment effectiveness for such a largely prevalent condition as musculoskeletal disorders, which represent the first indication to physiotherapy in the world. As a result, our findings may directly impact on important issues, such as the delivery of optimal care to patients and the liability of healthcare providers. A better adherence to the most rigorous scientific methods is imperative to improve the quality of scientific research and, thus, its impact on clinical practice.

**What was already known on this topic:** Systematic reviews have a fundamental role in summarising evidence on a given topic, thus providing guidance for best practices and guideline development. However, the methodological quality of these SR-MA on musculoskeletal conditions is uncertain.

**What this study adds:** Over the past ten years, nine out of ten systematic reviews are of critically low quality; this directly impacts the Evidence-Based Practice of physiotherapists and brings the urgency of improving the quality of secondary literature. The h-index of the last author is the only predictor of quality in our sample of 100 SRs.

## Data Availability

All data relevant to the study are included in the article or uploaded as supplementary information.

## FOOTNOTES

## Acknowledgments

We would like to thank Dr Beverly Shea for her feedback on some AMSTAR-2 details.

## CRediT author statement

Conceptualisation: all authors; methodology: NF, MDB, GC; investigation: NF, ER, AB; data curation: NF, ER, AB, GC; formal analysis: GC, MDB; supervision: PP, MDB; writing – original draft: NF, MDB; writing – review and editing: all authors.

^a^ EndNote Web, Clarivate, Philadelphia, USA

^b^ Microsoft Office Professional Plus 2021: Microsoft Excel. Microsoft.

^c^ StataCorp. 2021. Stata Statistical Software: Release 17. College Station, TX,USA: StataCorp LLC

