## Appendix 1 for "Quality of systematic reviews on physiotherapy interventions for musculoskeletal disorders is critically low: a meta-epidemiological study"

**MEDLINE (via Pubmed)**

| | |
|---|---|
| #7 | ((#1 OR #2 OR #3) AND (#4 OR #5) AND #6 |
| #6 | "systematic review"[Title/Abstract] |
| #5 | musculoskeletal[Title/Abstract] |
| #4 | musculoskeletal diseases[MeSH Terms] |
| #3 | modalities, physical therapy[MeSH Terms] |
| #2 | rehabilitation[Title/Abstract] |
| #1 | physiotherapy[Title/Abstract] |

**CINAHL (via EBSCOhost)**

| | |
|---|---|
| S9 | S6 AND S7 AND S8 |
| S8 | S4 OR S5 |
| S7 | S1 OR S2 OR S3 |
| S6 | AB systematic review |
| S5 | AB musculoskeletal |
| S4 | MH musculoskeletal diseases |
| S3 | MH physical therapy |
| S2 | AB rehabilitation |
| S1 | AB physiotherapy |

**Cochrane Database of Systematic Reviews**

| | |
|---|---|
| #6 | ((#1 OR #2 OR #3) AND (#4 OR #5) |
| #5 | (musculoskeletal):ti,ab,kw |
| #4 | MeSH descriptor: [Musculoskeletal Diseases] explode all trees |
| #3 | MeSH descriptor: [Physical Therapy Modalities] explode all trees |
| #2 | (rehabilitation):ti,ab,kw |
| #1 | (physiotherapy):ti,ab,kw |

**PEDro**

| | |
|---|---|
| Abstract & Title | musculoskeletal rehabilitation |
| Subdiscipline | musculoskeletal |
