## Appendix 2 for "Quality of systematic reviews on physiotherapy interventions for musculoskeletal disorders is critically low: a meta-epidemiological study"

| ID | First Author | Year | Title | Journal |
|---|---|---|---|---|
| 1 | Gutierrez Espinoza | 2022 | Effectiveness of manual therapy in patients with thumb carpometacarpal osteoarthritis: a systematic review and meta-analysis. | Physiotherapy Theory and Practice |
| 2 | Martimbianco | 2020 | Photobiomodulation with low-level laser therapy for treating Achilles tendinopathy: a systematic review and meta-analysis. | Clinical Rehabilitation |
| 3 | Ribeiro | 2018 | Laryngeal Manual Therapies for Behavioral Dysphonia: A Systematic Review and Meta-analysis. | Journal of voice |
| 4 | Varangot-Reille | 2022 | Effectiveness of Neural Mobilization Techniques in the Management of Musculoskeletal Neck Disorders with Nerve-Related Symptoms: A Systematic Review and Meta-Analysis with a Mapping Report. | Pain Medicine |
| 5 | Bertozzi | 2015 | Investigation of the effect of conservative interventions in thumb carpometacarpal osteoarthritis: systematic review and meta-analysis. | Disability and Rehabilitation |
| 6 | Tatsios | 2022 | The Effectiveness of Spinal, Diaphragmatic, and Specific Stabilization Exercise Manual Therapy and Respiratory-Related Interventions in Patients with Chronic Nonspecific Neck Pain: Systematic Review and Meta-Analysis. | Diagnostics |
| 7 | Zavala-Gonzalez | 2018 | The effectiveness of joint mobilization techniques for range of motion in adult patients with primary adhesive capsulitis of the shoulder: a systematic review and meta-analysis. | Medwave |
| 8 | Ryan | 2021 | Effect of Action Observation Therapy in the Rehabilitation of Neurologic and Musculoskeletal Conditions: A Systematic Review. | Archives of Rehabilitation Research and Clinical Translation |
| 9 | Ahern | 2018 | The effectiveness of physical therapies for patients with base of thumb osteoarthritis: Systematic review and meta-analysis. | Musculoskeletal Science and Practice |
| 10 | Tanaka | 2013 | Efficacy of strengthening or aerobic exercise on pain relief in people with knee osteoarthritis: a systematic review and meta-analysis of randomized controlled trials. | Clinical Rehabilitation |
| 11 | Beumer | 2016 | Effects of exercise and manual therapy on pain associated with hip osteoarthritis: a systematic review and meta-analysis. | British Journal of Sports Medicine |
| 12 | Kamonseki | 2021 | Effects of manual therapy on fear avoidance, kinesiophobia and pain catastrophizing in individuals with chronic musculoskeletal pain: Systematic review and meta-analysis. | Musculoskeletal Science and Practice |
| 13 | Li | 2013 | Electromagnetic fields for treating osteoarthritis | Cochrane Database of Systematic Reviews |
| 14 | Marin | 2017 | Multidisciplinary biopsychosocial rehabilitation for subacute low back pain | Cochrane Database of Systematic Reviews |
| 15 | Anwer | 2015 | Effects of Exercise on Spinal Deformities and Quality of Life in Patients with Adolescent Idiopathic Scoliosis. | BioMed Research International |
| 16 | Shi | 2018 | Aquatic Exercises in the Treatment of Low Back Pain: A Systematic Review of the Literature and Meta-Analysis of Eight Studies. | American Journal of Physical Medicine and Rehabilitation |
| 17 | Saracoglu | 2022 | Efficacy of adding pain neuroscience education to a multimodal treatment in fibromyalgia: A systematic review and meta-analysis. | International Journal of Rheumatic Diseases |
| 18 | Vallandingham | 2019 | Changes in Dorsiflexion and Dynamic Postural Control After Mobilizations in Individuals With Chronic Ankle Instability: A Systematic Review and Meta-Analysis. | Journal of Athletic Training |
| 19 | Cheng | 2019 | Effectiveness of Tai Chi on fibromyalgia patients: A meta-analysis of randomized controlled trials. | Complementary therapies in medicine |

| | | | | |
|---|---|---|---|---|
| 20 | Murphy | 2019 | Efficacy of heavy eccentric calf training for treating mid-portion Achilles tendinopathy: a systematic review and meta-analysis. | British Journal of Sports Medicine |
| 21 | Molina-Alvarez | 2022 | Manual Therapy Effect in Placebo-Controlled Trials: A Systematic Review and Meta-Analysis. | International Journal of Environmental Research and Public Health |
| 22 | Arora | 2022 | Physical modalities with eccentric exercise are no better than eccentric exercise alone in the treatment of chronic achilles tendinopathy: A systematic review and meta-analysis. | The Foot |
| 23 | Zhang | 2017 | The Effects of Traditional Chinese Exercise in Treating Knee Osteoarthritis: A Systematic Review and Meta-Analysis. | PLoS One |
| 24 | Nahon | 2021 | Physical therapy interventions for the treatment of delayed onset muscle soreness (DOMS): Systematic review and meta-analysis. | Physical Therapy in Sport |
| 25 | Fernandez | 2016 | Surgery or physical activity in the management of sciatica: a systematic review and meta-analysis. | European Spine Journal |
| 26 | Nicolson | 2017 | Interventions to increase adherence to therapeutic exercise in older adults with low back pain and/or hip/knee osteoarthritis: a systematic review and meta-analysis. | British Journal of Sports Medicine |
| 27 | Salazar | 2017 | Electric Stimulation for Pain Relief in Patients with Fibromyalgia: A Systematic Review and Meta-analysis of Randomized Controlled Trials. | Pain Physician |
| 28 | Li | 2022 | Effectiveness of motor imagery for improving functional performance after total knee arthroplasty: a systematic review with meta-analysis. | Journal of Orthopaedic Surgery and Research |
| 29 | MollàCasanova | 2021 | Effects of balance training on functionality, ankle instability, and dynamic balance outcomes in people with chronic ankle instability: Systematic review and meta-analysis. | Clinical Rehabilitation |
| 30 | Chen | 2016 | The effect of Tai Chi on four chronic conditions-cancer, osteoarthritis, heart failure and chronic obstructive pulmonary disease: a systematic review and meta-analyses. | British Journal of Sports Medicine |
| 31 | Saragiotto | 2020 | The Effectiveness of Strategies to Promote Walking in People With Musculoskeletal Disorders: A Systematic Review With Meta-analysis. | Journal of Orthopaedic and Sports Physical Therapy |
| 32 | Tamaoki | 2019 | Surgical versus conservative interventions for treating acromioclavicular dislocation of the shoulder in adults | Cochrane Database of Systematic Reviews |
| 33 | Tamin | 2018 | Exercise Intervention for Chronic Pain Management, Muscle Strengthening, and Functional Score in Obese Patients with Chronic Musculoskeletal Pain: A Systematic Review and Meta-analysis. | Acta Medica Indonesiana |
| 34 | Bini | 2022 | The effectiveness of manual and exercise therapy on headache intensity and frequency among patients with cervicogenic headache: a systematic review and meta-analysis. | Chiropractic & Manual Therapies |
| 35 | Buhagiar | 2019 | Assessment of Outcomes of Inpatient or Clinic-Based vs Home-Based Rehabilitation After Total Knee Arthroplasty: A Systematic Review and Meta-analysis. | JAMA Network Open |
| 36 | Heywood | 2017 | Effectiveness of Aquatic Exercise in Improving Lower Limb Strength in Musculoskeletal Conditions: A Systematic Review and Meta-Analysis. | Archives of Physical Medicine and Rehabilitation |
| 37 | Liu | 2018 | Evidence for Dry Needling in the Management of Myofascial Trigger Points Associated With Low Back Pain: A Systematic Review and Meta-Analysis. | Archives of Physical Medicine and Rehabilitation |
| 38 | Guzman-Pavon | 2020 | Effect of Physical Exercise Programs on Myofascial Trigger Points-Related Dysfunctions: A Systematic Review and Meta-analysis. | Pain Medicine |
| 39 | Burger | 2018 | The effectiveness of proprioceptive and neuromuscular training compared to bracing in reducing the recurrence rate of ankle sprains in athletes: A systematic review and meta-analysis. | Journal of Back and Musculoskeletal Rehabilitation |

| | | | | |
|---|---|---|---|---|
| 40 | DosSantos | 2021 | The effects of resistance training with blood flow restriction on muscle strength, muscle hypertrophy and functionality in patients with osteoarthritis and rheumatoid arthritis: A systematic review with meta-analysis. | PLOS ONE |
| 41 | Williams | 2018 | Exercise for rheumatoid arthritis of the hand | Cochrane Database of Systematic Reviews |
| 42 | Zou | 2020 | Traditional Chinese Eight Brocade Exercise Prescription for Ankylosing Spondylitis: A Quantitative Synthesis. | Complementary Medicine Research |
| 43 | Lin | 2020 | Comparative effects of combined physical therapy with Kinesio taping and physical therapy in patients with knee osteoarthritis: a systematic review and meta-analysis. | Clinical Rehabilitation |
| 44 | Liao | 2019 | Clinical efficacy of extracorporeal shockwave therapy for knee osteoarthritis: a systematic review and meta-regression of randomized controlled trials. | Clinical Rehabilitation |
| 45 | Ye | 2020 | Effectiveness of Elastic Taping in Patients With Knee Osteoarthritis: A Systematic Review and Meta-Analysis. | American Journal of Physical Medicine & Rehabilitation |
| 46 | Hamada | 2022 | The effectiveness of group education in people over 50 years old with knee pain: A systematic review and meta-analysis of randomized control trials. | Musculoskeletal Science and Practice |
| 47 | Lafrance | 2021 | Motor Control Exercises Compared to Strengthening Exercises for Upper- and Lower-Extremity Musculoskeletal Disorders: A Systematic Review With Meta-Analyses of Randomized Controlled Trials. | Physical Therapy |
| 48 | Ong | 2014 | The effect of dry needling for myofascial trigger points in the neck and shoulders: a systematic review and meta-analysis. | Journal of Bodywork & Movement Therapies |
| 49 | Cochrane | 2017 | Early interventions to promote work participation in people with regional musculoskeletal pain: a systematic review and meta-analysis. | Clinical Rehabilitation |
| 50 | Bartels | 2016 | Aquatic exercise for the treatment of knee and hip osteoarthritis | Cochrane Database of Systematic Reviews |
| 51 | Castro | 2021 | Effectiveness of conservative therapy in tendinopathy-related shoulder pain: A systematic review of randomized controlled trials. | Physical Therapy in Sport |
| 52 | Rogan | 2019 | Effects of Hip Abductor Muscles Exercises on Pain and Function in Patients With Patellofemoral Pain: A Systematic Review and Meta-Analysis. | Journal of Strength and Conditioning Research |
| 53 | Sieczkowska | 2019 | Effect of yoga on the quality of life of patients with rheumatic diseases: Systematic review with meta-analysis. | Complementary Therapies in Medicine |
| 54 | Luan | 2021 | Does Strength Training for Chronic Ankle Instability Improve Balance and Patient-Reported Outcomes and by Clinically Detectable Amounts? A Systematic Review and Meta-Analysis. | Physical Therapy |
| 55 | Thompson | 2019 | Effectiveness of scoliosis-specific exercises for adolescent idiopathic scoliosis compared with other non-surgical interventions: a systematic review and meta-analysis. | Physiotherapy |
| 56 | Webb | 2016 | Myofascial techniques: What are their effects on joint range of motion and pain? - A systematic review and meta-analysis of randomised controlled trials. | Journal of Bodywork & Movement Therapies |
| 57 | Alrawashdeh | 2021 | Effectiveness of total knee arthroplasty rehabilitation programmes: A systematic review and meta-analysis. | Journal of Rehabilitation Medicine |
| 58 | Ouellet | 2021 | Region-specific Exercises vs General Exercises in the Management of Spinal and Peripheral Musculoskeletal Disorders: A Systematic Review With Meta-analyses of Randomized Controlled Trials. | Archives of Physical Medicine and Rehabilitation |

| | | | | |
|---|---|---|---|---|
| 59 | Nazari | 2019 | The effectiveness of surgical vs conservative interventions on pain and function in patients with shoulder impingement syndrome. A systematic review and meta-analysis. | PLOS ONE |
| 60 | Albuquerque | 2022 | Effects of different protocols of physical exercise on fibromyalgia syndrome treatment: systematic review and meta-analysis of randomized controlled trials. | Rheumatology International |
| 61 | Ma | 2020 | The efficacy and safety of extracorporeal shockwave therapy in knee osteoarthritis: A systematic review and meta-analysis. | International Journal of Surgery |
| 62 | Martins | 2016 | Efficacy of musculoskeletal manual approach in the treatment of temporomandibular joint disorder: A systematic review with meta-analysis. | Manual Therapy |
| 63 | Ferreira | 2019 | Non-Pharmacological and Non-Surgical Interventions for Knee Osteoarthritis: A Systematic Review and Meta-Analysis. | Acta Reumatologica Portuguesa |
| 64 | Husted | 2020 | The relationship between prescribed pre-operative knee-extensor exercise dosage and effect on knee-extensor strength prior to and following total knee arthroplasty: a systematic review and meta-regression analysis of randomized controlled trials. | Osteoarthritis and Cartilage |
| 65 | Frutiger | 2021 | Systematic Review and Meta-Analysis Suggest Strength Training and Workplace Modifications May Reduce Neck Pain in Office Workers. | Pain Practice |
| 66 | Chen | 2016 | Transcutaneous Electrical Nerve Stimulation in Patients With Knee Osteoarthritis: Evidence From Randomized-controlled Trials. | The Clinical Journal of Pain |
| 67 | Flores | 2018 | Exercise training for improving outcomes post-burns: a systematic review and meta-analysis. | Clinical Rehabilitation |
| 68 | SosaReina | 2017 | Effectiveness of Therapeutic Exercise in Fibromyalgia Syndrome: A Systematic Review and Meta-Analysis of Randomized Clinical Trials. | BioMed research international. |
| 69 | Umehara | 2018 | Effective exercise intervention period for improving body function or activity in patients with knee osteoarthritis undergoing total knee arthroplasty: a systematic review and meta-analysis. | Brazilian journal of physical therapy. |
| 70 | CadellansArroniz | 2021 | The effectiveness of diacutaneous fibrolysis on pain, range of motion and functionality in musculoskeletal disorders: A systematic review and meta-analysis. | Clinical rehabilitation |
| 71 | Celik | 2020 | The clinical efficacy of kinesio taping in shoulder disorders: a systematic review and meta analysis. | Clinical rehabilitation. |
| 72 | Bizzarri | 2018 | Thoracic manual therapy is not more effective than placebo thoracic manual therapy in patients with shoulder dysfunctions: A systematic review with meta-analysis. | Musculoskeletal science & practice |
| 73 | Liu | 2015 | Effectiveness of dry needling for myofascial trigger points associated with neck and shoulder pain: a systematic review and meta-analysis. | Archives of Physical Medicine and Rehabilitation |
| 74 | Fellas | 2017 | Physical and Mechanical Therapies for Lower-Limb Problems in Juvenile Idiopathic Arthritis(A Systematic Review with Meta-Analysis). | Journal of the American Podiatric Medical Association |
| 75 | Salamh | 2017 | Treatment effectiveness and fidelity of manual therapy to the knee: A systematic review and meta-analysis. | Musculoskeletal care |
| 76 | GarciaHermoso | 2015 | Effects of exercise on functional aerobic capacity in adults with fibromyalgia syndrome: A systematic review of randomized controlled trials. | Journal of back and musculoskeletal rehabilitation |
| 77 | Tanaka | 2016 | Effects of exercise therapy on walking ability in individuals with knee osteoarthritis: a systematic review and meta-analysis of randomised controlled trials. | Clinical rehabilitation. |
| 78 | Cudejko | 2018 | Effect of Soft Braces on Pain and Physical Function in Patients With Knee Osteoarthritis: Systematic Review With Meta-Analyses. | Archives of Physical Medicine and Rehabilitation |
| 79 | Yin | 2022 | The effect of vibration training on delayed muscle soreness: A meta-analysis. | Medicine |

| | | | | |
|---|---|---|---|---|
| 80 | ArribasRomano | 2020 | Efficacy of Physical Therapy on Nociceptive Pain Processing Alterations in Patients with Chronic Musculoskeletal Pain: A Systematic Review and Meta-analysis. | Pain Medicine (United States) |
| 81 | Ammendolia | 2013 | Nonoperative treatment for lumbar spinal stenosis with neurogenic claudication | Cochrane Database of Systematic Reviews |
| 82 | Wu | 2022 | Benefits of Exergame Training for Female Patients With Fibromyalgia: A Systematic Review and Meta-Analysis of Randomized Controlled Trials. | Archives of Physical Medicine and Rehabilitation |
| 83 | Bartholdy | 2017 | The role of muscle strengthening in exercise therapy for knee osteoarthritis: A systematic review and meta-regression analysis of randomized trials. | Seminars in Arthritis and Rheumatism |
| 84 | Fransen | 2015 | Exercise for osteoarthritis of the knee | Cochrane Database of Systematic Reviews |
| 85 | VanGinckel | 2019 | Effects of long-term exercise therapy on knee joint structure in people with knee osteoarthritis: A systematic review and meta-analysis. | Seminars in Arthritis and Rheumatism |
| 86 | Waller | 2014 | Effect of therapeutic aquatic exercise on symptoms and function associated with lower limb osteoarthritis: systematic review with meta-analysis. | Physical therapy |
| 87 | Rubinstein | 2019 | Benefits and harms of spinal manipulative therapy for the treatment of chronic low back pain: systematic review and meta-analysis of randomised controlled trials. | BMJ : British medical journal / British Medical Association |
| 88 | vanDenDolder | 2014 | Effectiveness of soft tissue massage and exercise for the treatment of non-specific shoulder pain: a systematic review with meta-analysis. | British journal of sports medicine. |
| 89 | McGregor | 2013 | Rehabilitation following surgery for lumbar spinal stenosis | Cochrane Database of Systematic Reviews |
| 90 | Harpham | 2022 | The effect of exercise training programs with aerobic components on C-reactive protein, erythrocyte sedimentation rate and self-assessed disease activity in people with ankylosing spondylitis: A systematic review and meta-analysis. | International journal of rheumatic diseases |
| 91 | Sieczkowska | 2020 | Effects of resistance training on the health-related quality of life of patients with rheumatic diseases: Systematic review with meta-analysis and meta-regression. | Seminars in arthritis and rheumatism |
| 92 | Kroon | 2014 | Self-management education programmes for osteoarthritis | Cochrane Database of Systematic Reviews |
| 93 | Stania | 2022 | Treatment of Jumper's Knee with Extracorporeal Shockwave Therapy: A Systematic Review and Meta-Analysis. | Journal of human kinetics |
| 94 | Liao | 2020 | Effects of Muscle Strength Training on Muscle Mass Gain and Hypertrophy in Older Adults With Osteoarthritis: A Systematic Review and Meta-Analysis. | Arthritis Care & Research |
| 95 | Luo | 2017 | The effect of whole-body vibration therapy on bone metabolism, motor function, and anthropometric parameters in women with postmenopausal osteoporosis. | Disability and rehabilitation |
| 96 | Anwer | 2016 | Effect of whole body vibration training on quadriceps muscle strength in individuals with knee osteoarthritis: a systematic review and meta-analysis. | Physiotherapy |
| 97 | Runge | 2022 | The Benefits of Adding Manual Therapy to Exercise Therapy for Improving Pain and Function in Patients With Knee or Hip Osteoarthritis: A Systematic Review With Meta-analysis. | journal of orthopaedic & sports physical therapy |
| 98 | Machado | 2016 | Can Water Temperature and Immersion Time Influence the Effect of Cold Water Immersion on Muscle Soreness? A Systematic Review and Meta-Analysis. | Sports Medicine |
| 99 | Osteras | 2017 | Exercise for hand osteoarthritis | Cochrane Database of Systematic Reviews |

| | | | | |
|---|---|---|---|---|
| 100 | Quentin | 2021 | Effect of Home Exercise Training in Patients with Nonspecific Low-Back Pain: A Systematic Review and Meta-Analysis. | International Journal of Environmental Research and Public Health |
