## Appendix 3 for "Quality of systematic reviews on physiotherapy interventions for musculoskeletal disorders is critically low: a meta-epidemiological study"

**List of studies excluded with reasons**

**Wrong study design**

Alex, James L. N. er, Adam G. Culvenor, Richard R. T. Johnston, Allison M. Ezzat, and Christian J. Barton. 'Strategies to Prevent and Manage Running-Related Knee Injuries: A Systematic Review of Randomised Controlled Trials.' British Journal of Sports Medicine 56, no. 22 (2022): 1307–19. https://doi.org/10.1136/bjsports-2022-105553.

Almeida, MO, BNG Silva, RB Andriolo, ÁN Atallah, and MS Peccin. 'Conservative Interventions for Treating Exercise-related Musculotendinous, Ligamentous and Osseous Groin Pain'. Cochrane Database of Systematic Reviews, no. 6 (2013). https://doi.org/10.1002/14651858.CD009565.pub2.

Ammendolia, Carlo, Corey Hofkirchner, Joshua Plener, André Bussières, Michael J. Schneider, James J. Young, Andrea D. Furlan, et al. 'Non-Operative Treatment for Lumbar Spinal Stenosis with Neurogenic Claudication: An Updated Systematic Review.' BMJ Open 12, no. 1 (2022): e057724. https://doi.org/10.1136/bmjopen-2021-057724.

Barton, Christian, Balach, Vivek ar, Simon Lack, and Dylan Morrissey. 'Patellar Taping for Patellofemoral Pain: A Systematic Review and Meta-Analysis to Evaluate Clinical Outcomes and Biomechanical Mechanisms.' British Journal of Sports Medicine 48, no. 6 (2014): 417–24. https://doi.org/10.1136/bjsports-2013-092437.

Bexkens, Rens, Frederic J. Washburn, Denise Eygendaal, Michel P. J. van den Bekerom, and Luke S. Oh. 'Effectiveness of Reduction Maneuvers in the Treatment of Nursemaid's Elbow: A Systematic Review and Meta-Analysis.' The American Journal of Emergency Medicine 35, no. 1 (2017): 159–63. https://doi.org/10.1016/j.ajem.2016.10.059.

Bina, S, V Pacey, EH Barnes, J Burns, and K Gray. 'Interventions for Congenital Talipes Equinovarus (Clubfoot)'. Cochrane Database of Systematic Reviews, no. 5 (2020). https://doi.org/10.1002/14651858.CD008602.pub4.

Challoumas, Dimitris, Mairiosa Biddle, Michael McLean, and Neal L. Millar. 'Comparison of Treatments for Frozen Shoulder: A Systematic Review and Meta-Analysis.' JAMA Network Open 3, no. 12 (2020): e2029581. https://doi.org/10.1001/jamanetworkopen.2020.29581.

Chen, Zhiqing, and Nancy A. Baker. 'Effectiveness of Eccentric Strengthening in the Treatment of Lateral Elbow Tendinopathy: A Systematic Review with Meta-Analysis.' Journal of Hand Therapy: Official Journal of the American Society of Hand Therapists 34, no. 1 (2021): 18–28. https://doi.org/10.1016/j.jht.2020.02.002.

Cohen, Dan, NhatChinh Le, Alex Zakharia, er, Benjamin Blackman, and Darren de Sa. 'MPFL Reconstruction Results in Lower Redislocation Rates and Higher Functional Outcomes than Rehabilitation: A Systematic Review and Meta-Analysis.' Knee Surgery, Sports Traumatology, Arthroscopy: Official Journal of the ESSKA 30, no. 11 (2022): 3784–95. https://doi.org/10.1007/s00167-022-07003-5.

Conley, Caitlin E. W., Carl G. Mattacola, Kate N. Jochimsen, Emily V. Dressler, Christian Lattermann, and Jennifer S. Howard. 'A Comparison of Neuromuscular Electrical Stimulation Parameters for Postoperative Quadriceps Strength in Patients After Knee Surgery: A Systematic Review.' Sports Health 13, no. 2 (2021): 116–27. https://doi.org/10.1177/1941738120964817.

Ezzat, Allison M., Katie MacPherson, Jenny Leese, and Linda C. Li. 'The Effects of Interventions to Increase Exercise Adherence in People with Arthritis: A Systematic Review.' Musculoskeletal Care 13, no. 1 (2015): 1–18. https://doi.org/10.1002/msc.1084.

Ezzati, Kamran, E.-Liisa Laakso, Amir Salari, Anahita Hasannejad, Reza Fekrazad, and Arash Aris. 'The Beneficial Effects of High-Intensity Laser Therapy and Co-Interventions on Musculoskeletal Pain Management: A Systematic Review.' Journal of Lasers in Medical Sciences 11, no. 1 (2020): 81–90. https://doi.org/10.15171/jlms.2020.14.

Feger, Mark A., John Goetschius, Hailey Love, Sue A. Saliba, and Jay Hertel. 'Electrical Stimulation as a Treatment Intervention to Improve Function, Edema or Pain Following Acute Lateral Ankle Sprains: A Systematic Review.' Physical Therapy in Sport: Official Journal of the Association of Chartered Physiotherapists in Sports Medicine 16, no. 4 (2015): 361–69. https://doi.org/10.1016/j.ptsp.2015.01.001.

Filbay, S. R., A. G. Culvenor, I. N. Ackerman, T. G. Russell, and K. M. Crossley. 'Quality of Life in Anterior Cruciate Ligament-Deficient Individuals: A Systematic Review and Meta-Analysis.' British Journal of Sports Medicine 49, no. 16 (2015): 1033–41. https://doi.org/10.1136/bjsports-2015-094864.

Fong Yan, Alycia, Stephen Cobley, Cliffton Chan, Evangelos Pappas, Leslie L. Nicholson, Rachel E. Ward, Roslyn E. Murdoch, et al. 'The Effectiveness of Dance Interventions on Physical Health Outcomes Compared to Other Forms of Physical Activity: A Systematic Review and Meta-Analysis.' Sports Medicine (Auckland, N.Z.) 48, no. 4 (2018): 933–51. https://doi.org/10.1007/s40279-017-0853-5.

Gava, V, er, Larissa Pechincha Ribeiro, Rodrigo Py Gonçalves Barreto, and Paula Rezende Camargo. 'Effectiveness of Physical Therapy given by Telerehabilitation on Pain and Disability of Individuals with Shoulder Pain: A Systematic Review.' Clinical Rehabilitation 36, no. 6 (2022): 715–25. https://doi.org/10.1177/02692155221083496.

Gluppe, S, ra, Marie Ellström Engh, and Kari Bø. 'What Is the Evidence for Abdominal and Pelvic Floor Muscle Training to Treat Diastasis Recti Abdominis Postpartum? A Systematic Review with Meta-Analysis.' Brazilian Journal of Physical Therapy 25, no. 6 (2021): 664–75. https://doi.org/10.1016/j.bjpt.2021.06.006.

Gumaa, Mohammed, and Aliaa Rehan Youssef. 'Is Virtual Reality Effective in Orthopedic Rehabilitation? A Systematic Review and Meta-Analysis.' Physical Therapy 99, no. 10 (2019): 1304–25. https://doi.org/10.1093/ptj/pzz093.

Gwinnutt JM, Wieczorek M, Cavalli G, Balanescu A, Bischoff-Ferrari HA, Boonen A, de Souza S, et al. 'Effects of Physical Exercise and Body Weight on Disease-Specific Outcomes of People with Rheumatic and Musculoskeletal Diseases (RMDs): Systematic Reviews and Meta-Analyses Informing the 2021 EULAR Recommendations for Lifestyle Improvements in People with RMDs [with Consumer Summary]'. RMD Open 2022 Mar;8(1): E002168, 2022.

Haik, M. N., F. Alburquerque-Sendín, R. F. C. Moreira, E. D. Pires, and P. R. Camargo. 'Effectiveness of Physical Therapy Treatment of Clearly Defined Subacromial Pain: A Systematic Review of Randomised Controlled Trials.' British Journal of Sports Medicine 50, no. 18 (2016): 1124–34. https://doi.org/10.1136/bjsports-2015-095771.

Hall, M., F. Dobson, M. Plinsinga, C. Mailloux, S. Starkey, E. Smits, P. Hodges, B. Vicenzino, S. M. Schabrun, and H. Masse-Alarie. 'Effect of Exercise on Pain Processing and Motor Output in People with Knee Osteoarthritis: A Systematic Review and Meta-Analysis.' Osteoarthritis and Cartilage 28, no. 12 (2020): 1501–13. https://doi.org/10.1016/j.joca.2020.07.009.

Häußer, Jonathan, Juliane Wieber, and Philip Catalá-Lehnen. 'The Use of Extracorporeal Shock Wave Therapy for the Treatment of Bone Marrow Oedema - a Systematic Review and Meta-Analysis.' Journal of Orthopaedic Surgery and Research 16, no. 1 (2021): 369. https://doi.org/10.1186/s13018-021-02484-5.

Henriksen, Marius, Julie B. Hansen, Louise Klokker, Henning Bliddal, and Robin Christensen. 'Comparable Effects of Exercise and Analgesics for Pain Secondary to Knee Osteoarthritis: A Meta-Analysis of Trials Included in Cochrane Systematic Reviews.' Journal of Comparative Effectiveness Research 5, no. 4 (2016): 417–31. https://doi.org/10.2217/cer-2016-0007.

Higgins, Trevor R., David A. Greene, and Michael K. Baker. 'Effects of Cold Water Immersion and Contrast Water Therapy for Recovery From Team Sport: A Systematic Review and Meta-Analysis.' Journal of Strength and Conditioning Research 31, no. 5 (2017): 1443–60. https://doi.org/10.1519/JSC.0000000000001559.

Iijima, Hirotaka, and Masaki Takahashi. 'Microcurrent Therapy as a Therapeutic Modality for Musculoskeletal Pain: A Systematic Review Accelerating the Translation From Clinical Trials to Patient Care.' Archives of Rehabilitation Research and Clinical Translation 3, no. 3 (2021): 100145. https://doi.org/10.1016/j.arrct.2021.100145.

Johnson, Mark I., Leica S. Claydon, G. Peter Herbison, Gareth Jones, and Carole A. Paley. 'Transcutaneous Electrical Nerve Stimulation (TENS) for Fibromyalgia in Adults.' The Cochrane Database of Systematic Reviews 10, no. 10 (2017): CD012172. https://doi.org/10.1002/14651858.CD012172.pub2.

Kelley, George A., and Kristi S. Kelley. 'Effects of Exercise on Depressive Symptoms in Adults with Arthritis and Other Rheumatic Disease: A Systematic Review of Meta-Analyses.' BMC Musculoskeletal Disorders 15 (2014): 121. https://doi.org/10.1186/1471-2474-15-121.

Kosik, Kyle B., Ryan S. McCann, Masafumi Terada, and Phillip A. Gribble. 'Therapeutic Interventions for Improving Self-Reported Function in Patients with Chronic Ankle Instability: A Systematic Review.' British Journal of Sports Medicine 51, no. 2 (2017): 105–12. https://doi.org/10.1136/bjsports-2016-096534.

Krul, M, JC van der Wouden, EJ Kruithof, LWA van Suijlekom-Smit, and BW Koes. 'Manipulative Interventions for Reducing Pulled Elbow in Young Children'. Cochrane Database of Systematic Reviews, no. 7 (2017). https://doi.org/10.1002/14651858.CD007759.pub4.

Lee, O.-Sung, Soyeon Ahn, and Yong Seuk Lee. 'Effect and Safety of Early Weight-Bearing on the Outcome after Open-Wedge High Tibial Osteotomy: A Systematic Review and Meta-Analysis.' Archives of Orthopaedic and Trauma Surgery 137, no. 7 (2017): 903–11. https://doi.org/10.1007/s00402-017-2703-1.

Li, Shuoqi, Wei Hui Ng, Sumayeh Abujaber, and Shazlin Shaharudin. 'Effects of Resistance Training on Gait Velocity and Knee Adduction Moment in Knee Osteoarthritis Patients: A Systematic Review and Meta-Analysis.' Scientific Reports 11, no. 1 (2021): 16104. https://doi.org/10.1038/s41598-021-95426-4.

Loew, LM, L Brosseau, P Tugwell, GA Wells, V Welch, B Shea, S Poitras, G De Angelis, and P Rahman. 'Deep Transverse Friction Massage for Treating Lateral Elbow or Lateral Knee Tendinitis'. Cochrane Database of Systematic Reviews, no. 11 (2014). https://doi.org/10.1002/14651858.CD003528.pub2.

Logan, Catherine A., Abhiram R. Bhashyam, Ashley J. Tisosky, Daniel B. Haber, Anna Jorgensen, Adam Roy, and Matthew T. Provencher. 'Systematic Review of the Effect of Taping Techniques on Patellofemoral Pain Syndrome.' Sports Health 9, no. 5 (2017): 456–61. https://doi.org/10.1177/1941738117710938.

Lomas-Vega, Rafael, María Victoria Garrido-Jaut, Alma Rus, and Rafael Del-Pino-Casado. 'Effectiveness of Global Postural Re-Education for Treatment of Spinal Disorders: A Meta-Analysis.' American Journal of Physical Medicine & Rehabilitation 96, no. 2 (2017): 124–30. https://doi.org/10.1097/PHM.0000000000000575.

Longo, Umile Giuseppe, Just A. van der Linde, Mattia Loppini, Vito Coco, Rudolf W. Poolman, and Vincenzo Denaro. 'Surgical Versus Nonoperative Treatment in Patients Up to 18 Years Old With Traumatic Shoulder Instability: A Systematic Review and Quantitative Synthesis of the Literature.' Arthroscopy: The Journal of Arthroscopic & Related Surgery: Official Publication of the Arthroscopy Association of North America and the International Arthroscopy Association 32, no. 5 (2016): 944–52. https://doi.org/10.1016/j.arthro.2015.10.020.

Martimbianco, ALC, GJM Porfírio, RL Pacheco, MR Torloni, and R Riera. 'Transcutaneous Electrical Nerve Stimulation (TENS) for Chronic Neck Pain'. Cochrane Database of Systematic Reviews, no. 12 (2019). https://doi.org/10.1002/14651858.CD011927.pub2.

Meer, Hedwig A. van der, Leticia B. Calixtre, Raoul H. H. Engelbert, Corine M. Visscher, Nijhuis-van der S, Maria Wg en, and Caroline M. Speksnijder. 'Effects of Physical Therapy for Temporomandibular Disorders on Headache Pain Intensity: A Systematic Review.' Musculoskeletal Science & Practice 50 (2020): 102277. https://doi.org/10.1016/j.msksp.2020.102277.

Mei, Jin, Lili Pang, and Zhongchao Jiang. 'The Effect of Extracorporeal Shock Wave on Osteonecrosis of Femoral Head: A Systematic Review and Meta-Analysis.' The Physician and Sportsmedicine 50, no. 4 (2022): 280–88. https://doi.org/10.1080/00913847.2021.1936685.

Merza, Eman, Stephen Pearson, Glen Lichtwark, Meg Ollason, and Peter Malliaras. 'Immediate and Long-Term Effects of Mechanical Loading on Achilles Tendon Volume: A Systematic Review and Meta-Analysis.' Journal of Biomechanics 118 (2021): 110289. https://doi.org/10.1016/j.jbiomech.2021.110289.

Michaleff, Zoe A., and Steven J. Kamper. 'PEDro Systematic Review Update: The Effectiveness of Physiotherapy Exercises in Subacromical Impingement Syndrome.' British Journal of Sports Medicine 47, no. 14 (2013): 927–28. https://doi.org/10.1136/bjsports-2013-092750.

Monk, AP, LJ Davies, S Hopewell, K Harris, DJ Beard, and AJ Price. 'Surgical versus Conservative Interventions for Treating Anterior Cruciate Ligament Injuries'. Cochrane Database of Systematic Reviews, no. 4 (2016). https://doi.org/10.1002/14651858.CD011166.pub2.

Mudano, AS, P Tugwell, GA Wells, and JA Singh. 'Tai Chi for Rheumatoid Arthritis'. Cochrane Database of Systematic Reviews, no. 9 (2019). https://doi.org/10.1002/14651858.CD004849.pub2.

Murphy, Myles, Mervyn Travers, William Gibson, Paola Chivers, James Debenham, Sean Docking, and Ebonie Rio. 'Rate of Improvement of Pain and Function in Mid-Portion Achilles Tendinopathy with Loading Protocols: A Systematic Review and Longitudinal Meta-Analysis.' Sports Medicine (Auckland, N.Z.) 48, no. 8 (2018): 1875–91. https://doi.org/10.1007/s40279-018-0932-2.

Ojha, Heidi A., Nadia J. Wyrsta, Todd E. Davenport, William E. Egan, and Alfred C. Gellhorn. 'Timing of Physical Therapy Initiation for Nonsurgical Management of Musculoskeletal Disorders and Effects on Patient Outcomes: A Systematic Review.' The Journal of Orthopaedic and Sports Physical Therapy 46, no. 2 (2016): 56–70. https://doi.org/10.2519/jospt.2016.6138.

Page, MJ, S Green, S Kramer, RV Johnston, B McBain, and R Buchbinder. 'Electrotherapy Modalities for Adhesive Capsulitis (Frozen Shoulder)'. Cochrane Database of Systematic Reviews, no. 10 (2014). https://doi.org/10.1002/14651858.CD011324.

Page, MJ, S Green, B McBain, SJ Surace, J Deitch, N Lyttle, MA Mrocki, and R Buchbinder. 'Manual Therapy and Exercise for Rotator Cuff Disease'. Cochrane Database of Systematic Reviews, no. 6 (2016). https://doi.org/10.1002/14651858.CD012224.

Page, MJ, S Green, MA Mrocki, SJ Surace, J Deitch, B McBain, N Lyttle, and R Buchbinder. 'Electrotherapy Modalities for Rotator Cuff Disease'. Cochrane Database of Systematic Reviews, no. 6 (2016). https://doi.org/10.1002/14651858.CD012225.

Papaconstantinou, Efrosini, Carol Cancelliere, Leslie Verville, Jessica J. Wong, Gaelan Connell, Hainan Yu, Heather Shearer, et al. 'Effectiveness of Non-Pharmacological Interventions on Sleep Characteristics among Adults with Musculoskeletal Pain and a Comorbid Sleep Problem: A Systematic Review.' Chiropractic & Manual Therapies 29, no. 1 (2021): 23. https://doi.org/10.1186/s12998-021-00381-6.

Powden, Cameron J., Johanna M. Hoch, and Matthew C. Hoch. 'Rehabilitation and Improvement of Health-Related Quality-of-Life Detriments in Individuals With Chronic Ankle Instability: A Meta-Analysis.' Journal of Athletic Training 52, no. 8 (2017): 753–65. https://doi.org/10.4085/1062-6050-52.5.01.

Reyburn, Robert J., and Cameron J. Powden. 'Dynamic Balance Measures in Healthy and Chronic Ankle Instability Participants While Wearing Ankle Braces: Systematic Review With Meta-Analysis.' Journal of Sport Rehabilitation 30, no. 4 (2020): 660–67. https://doi.org/10.1123/jsr.2020-0224.

Salomon, Mattia, Chiara Pastore, Filippo Maselli, Mauro Di Bari, Raffaello Pellegrino, and Fabrizio Brindisino. 'Manipulation under Anesthesia versus Non-Surgical Treatment for Patients with Frozen Shoulder Contracture Syndrome: A Systematic Review.' International Journal of Environmental Research and Public Health 19, no. 15 (2022). https://doi.org/10.3390/ijerph19159715.

Saltychev, Mikhail, Rebecca A. Dutton, Katri Laimi, Gary S. Beaupré, Petri Virolainen, and Michael Fredericson. 'Effectiveness of Conservative Treatment for Patellofemoral Pain Syndrome: A Systematic Review and Meta-Analysis.' Journal of Rehabilitation Medicine 50, no. 5 (2018): 393–401. https://doi.org/10.2340/16501977-2295.

Sattler, Larissa Nicole, Wayne Anthony Hing, and Christopher John Vertullo. 'What Is the Evidence to Support Early Supervised Exercise Therapy after Primary Total Knee Replacement? A Systematic Review and Meta-Analysis.' BMC Musculoskeletal Disorders 20, no. 1 (2019): 42. https://doi.org/10.1186/s12891-019-2415-5.

Schubert, Ilona, Peter C. Strohm, Dirk Maier, and Jörn Zwingmann. 'Simple Traumatic Elbow Dislocations; Benefit from Early Functional Rehabilitation: A Systematic Review with Meta-Analysis Including PRISMA Criteria.' Medicine 100, no. 44 (2021): e27168. https://doi.org/10.1097/MD.0000000000027168.

Schulze, Nina B., Marianna de Melo Salemi, Geisa G. de Alencar, Marcela C. Moreira, and Gisela R. de Siqueira. 'Efficacy of Manual Therapy on Pain, Impact of Disease, and Quality of Life in the Treatment of Fibromyalgia: A Systematic Review.' Pain Physician 23, no. 5 (2020): 461–76.

Shekelle, Paul G., Ian A. Cook, Isomi M. Miake-Lye, Marika Suttorp Booth, Jessica M. Beroes, and Selene Mak. 'Benefits and Harms of Cranial Electrical Stimulation for Chronic Painful Conditions, Depression, Anxiety, and Insomnia: A Systematic Review.' Annals of Internal Medicine 168, no. 6 (2018): 414–21. https://doi.org/10.7326/M17-1970.

Sieczkowska, Sofia Mendes, Fabiana Infante Smaira, Bruna Caruso Mazzolani, Bruno Gualano, Hamilton Roschel, and Tiago Peçanha. 'Efficacy of Home-Based Physical Activity Interventions in Patients with Autoimmune Rheumatic Diseases: A Systematic Review and Meta-Analysis.' Seminars in Arthritis and Rheumatism 51, no. 3 (2021): 576–87. https://doi.org/10.1016/j.semarthrit.2021.04.004.

Smith, T. O., K. Postle, F. Penny, I. McNamara, and C. J. V. Mann. 'Is Reconstruction the Best Management Strategy for Anterior Cruciate Ligament Rupture? A Systematic Review and Meta-Analysis Comparing Anterior Cruciate Ligament Reconstruction versus Non-Operative Treatment.' The Knee 21, no. 2 (2014): 462–70. https://doi.org/10.1016/j.knee.2013.10.009.

Snowdon, Megan, and Casey L. Peiris. 'Physiotherapy Commenced Within the First Four Weeks Post-Spinal Surgery Is Safe and Effective: A Systematic Review and Meta-Analysis.' Archives of Physical Medicine and Rehabilitation 97, no. 2 (2016): 292–301. https://doi.org/10.1016/j.apmr.2015.09.003.

Stanhope J, Pisaniello D, and Weinstein P. 'The Effect of Strategies to Prevent and Manage Musicians' Musculoskeletal Symptoms: A Systematic Review'. Archives of Environmental & Occupational Health 2022;77(3):185-208, 2022.

Sturman, Sarah, and Clare Killingback. 'Is There a Dose Response Relationship between Soft Tissue Manual Therapy and Clinical Outcomes in Fibromyalgia?' Journal of Bodywork and Movement Therapies 24, no. 3 (2020): 141–53. https://doi.org/10.1016/j.jbmt.2020.02.010.

Su, Yanlin, Zhe Chen, and Wei Xie. 'Swimming as Treatment for Osteoporosis: A Systematic Review and Meta-Analysis.' BioMed Research International 2020 (2020): 6210201. https://doi.org/10.1155/2020/6210201.

Tedla, Jaya Shanker, and Devika Rani Sangadala. 'Proprioceptive Neuromuscular Facilitation Techniques in Adhesive Capsulitis: A Systematic Review and Meta-Analysis.' Journal of Musculoskeletal & Neuronal Interactions 19, no. 4 (2019): 482–91.

Teirlinck, Carolien H., Arianne P. Verhagen, Elja A. E. Reijneveld, Jos Runhaar, Marienke van Middelkoop, Leontien M. van Ravesteyn, Lotte Hermsen, Ingrid B. de Groot, and Sita M. A. Bierma-Zeinstra. 'Responders to Exercise Therapy in Patients with Osteoarthritis of the Hip: A Systematic Review and Meta-Analysis.' International Journal of Environmental Research and Public Health 17, no. 20 (2020). https://doi.org/10.3390/ijerph17207380.

Terada, Masafumi, Brian G. Pietrosimone, and Phillip A. Gribble. 'Therapeutic Interventions for Increasing Ankle Dorsiflexion after Ankle Sprain: A Systematic Review.' Journal of Athletic Training 48, no. 5 (2013): 696–709. https://doi.org/10.4085/1062-6050-48.4.11.

Vassão, Patricia Gabielli, Julia Parisi, Thaíse Fern Penha, a Campos, Ana Beatriz Balão, Ana Claudia Muniz Renno, and Mariana Arias Avila. 'Association of Photobiomodulation Therapy (PBMT) and Exercises Programs in Pain and Functional Capacity of Patients with Knee Osteoarthritis (KOA): A Systematic Review of Randomized Trials.' Lasers in Medical Science 36, no. 7 (2021): 1341–53. https://doi.org/10.1007/s10103-020-03223-8.

Verhagen, Arianne P., Sita M. A. Bierma-Zeinstra, Maarten Boers, Jefferson R. Cardoso, Johan Lambeck, Rob de Bie, and Henrica C. W. de Vet. 'Balneotherapy (or Spa Therapy) for Rheumatoid Arthritis.' The Cochrane Database of Systematic Reviews 2015, no. 4 (2015): CD000518. https://doi.org/10.1002/14651858.CD000518.pub2.

Winkler, S, ra L., Anthony E. Urbisci, and Thomas M. Best. 'Sustained Acoustic Medicine for the Treatment of Musculoskeletal Injuries: A Systematic Review and Meta-Analysis.' BMC Sports Science, Medicine & Rehabilitation 13, no. 1 (2021): 159. https://doi.org/10.1186/s13102-021-00383-0.

Young, Jodi L., Daniel I. Rhon, Rutger M. J. de Zoete, Clel, Joshua A., and Suzanne J. Snodgrass. 'The Influence of Dosing on Effect Size of Exercise Therapy for Musculoskeletal Foot and Ankle Disorders: A Systematic Review.' Brazilian Journal of Physical Therapy 22, no. 1 (2018): 20–32. https://doi.org/10.1016/j.bjpt.2017.10.001.

**Wrong population**

Antoniak, Anneka Elizabeth, and Carolyn A. Greig. 'The Effect of Combined Resistance Exercise Training and Vitamin D(3) Supplementation on Musculoskeletal Health and Function in Older Adults: A Systematic Review and Meta-Analysis.' BMJ Open 7, no. 7 (2017): e014619. https://doi.org/10.1136/bmjopen-2016-014619.

Berl, Rémi, Elena Marques-Sule, José Luis Marín-Mateo, Noemi Moreno-Segura, Ana López-Ridaura, Sent, and Trinidad reu-Mañó. 'Effects of the Feldenkrais Method as a Physiotherapy Tool: A Systematic Review and Meta-Analysis of Randomized Controlled Trials.' International Journal of Environmental Research and Public Health 19, no. 21 (2022). https://doi.org/10.3390/ijerph192113734.

Ghai, Shashank, Matthew Driller, and Ishan Ghai. 'Effects of Joint Stabilizers on Proprioception and Stability: A Systematic Review and Meta-Analysis.' Physical Therapy in Sport: Official Journal of the Association of Chartered Physiotherapists in Sports Medicine 25 (2017): 65–75. https://doi.org/10.1016/j.ptsp.2016.05.006.

**Wrong intervention**

Bouchard, Carl, Jean-Paul Goulet, Mehdi El-Ouazzani, and Alexis F. Turgeon. 'Temporomandibular Lavage Versus Nonsurgical Treatments for Temporomandibular Disorders: A Systematic Review and Meta-Analysis.' Journal of Oral and Maxillofacial Surgery: Official Journal of the American Association of Oral and Maxillofacial Surgeons 75, no. 7 (2017): 1352–62. https://doi.org/10.1016/j.joms.2016.12.027.

Oliveira Lima, Lívia de, Bruno T. Saragiotto, Leonardo Oliveira Pena Costa, Le Nogueira, ro Calazans, Ney Meziat-Filho, and Felipe J. J. Reis. 'Self-Guided Web-Based Pain Education for

People With Musculoskeletal Pain: A Systematic Review and Meta-Analysis.' Physical Therapy 101, no. 10 (2021): pzab167. https://doi.org/10.1093/ptj/pzab167.

Fan, Weiming, Yajian Wang, and Yu Zhao. 'Effects of Enhanced Recovery Rehabilitation Surgery Concepts on the Surgical Process, Postoperative Pain, Complications, and Prognosis of Discectomy in Patients with Lumbar Disc Herniation: A Systematic Review and Meta-Analysis.' Computational and Mathematical Methods in Medicine 2022 (2022): 9736470. https://doi.org/10.1155/2022/9736470.

Kessler, Christian S., Lea Pinders, Andreas Michalsen, and Holger Cramer. 'Ayurvedic Interventions for Osteoarthritis: A Systematic Review and Meta-Analysis.' Rheumatology International 35, no. 2 (2015): 211–32. https://doi.org/10.1007/s00296-014-3095-y.

Matthews, Barry G., Sheree E. Hurn, Michael P. Harding, Rachel A. Henry, and Robert S. Ware. 'The Effectiveness of Non-Surgical Interventions for Common Plantar Digital Compressive Neuropathy (Morton's Neuroma): A Systematic Review and Meta-Analysis.' Journal of Foot and Ankle Research 12 (2019): 12. https://doi.org/10.1186/s13047-019-0320-7.

Pushparaj, Hemkumar, Yasmine Hoydonckx, Nimish Mittal, Philip Peng, Steven P. Cohen, Xingshan Cao, and Anuj Bhatia. 'A Systematic Review and Meta-Analysis of Radiofrequency Procedures on Innervation to the Shoulder Joint for Relieving Chronic Pain.' European Journal of Pain (London, England) 25, no. 5 (2021): 986–1011. https://doi.org/10.1002/ejp.1735.

Vos, Lukas M., James J. R. Huddleston Slater, and Boudewijn Stegenga. 'Lavage Therapy versus Nonsurgical Therapy for the Treatment of Arthralgia of the Temporomandibular Joint: A Systematic Review of Randomized Controlled Trials.' Journal of Orofacial Pain 27, no. 2 (2013): 171–79. https://doi.org/10.11607/jop.1007.

**Wong publication type**

Fagundes, A. G., C. Fischer, C. E. Alves, and R. Poton. 'Effectiveness of Dry Needling Combined with a Treatment Programme for Musculoskeletal-Related Chronic Pain: A Systematic Review and Meta-Analysis.' International Journal of Sports Physical Therapy 14, no. 6 (2019): S18–19.
